# Gender Differences in the Diagnosis of Chronic Obstructive Pulmonary Disease after Spirometry

**DOI:** 10.1101/2024.07.18.24310648

**Authors:** Alexander T. Moffett, Scott D. Halpern, Gary E. Weissman

**Author notes:** **Corresponding Author:** Alexander T. Moffett, Hospital of the University of Pennsylvania, Division of Pulmonary, Allergy, and Critical Care Medicine, 3400 Spruce Street, Philadelphia, PA 19104. **Contributions:** All authors fulfill ICMJE criteria for authorship.

## Abstract

**Background:** Women are more likely than men to report delays in the diagnosis of chronic obstructive pulmonary disease (COPD), though the etiology of these delays is unknown. We sought to test whether delays in COPD diagnosis persist after the performance of spirometry.

**Methods:** We used the Optum Labs Data Warehouse to identify patients 18 years of age and older without a prior diagnosis of COPD, with a post-bronchodilator forced expiratory volume in 1 second (FEV_1_) to forced vital capacity (FVC) ratio of less than 0.7 on spirometry. We used a Cox proportional hazards model to compare the time to diagnosis after spirometry in men and women, adjusting for age, race, ethnicity, tobacco use, and post-bronchodilator FEV_1_/FVC.

**Results:** The probability of receiving a COPD diagnosis after the performance of spirometry was lower among women than men (adjusted hazard ratio [aHR] 0.66, 95% confidence interval [CI] 0.50 to 0.88)

**Conclusion:** In this retrospective cohort study of patients with spirometric evidence of obstruction, the time to diagnosis of COPD was greater among women than men. While previous vignette-based studies have found that gender differences in the diagnosis of COPD resolve with the performance of spirometry, we found that gender differences persist after spirometry has been performed. Clinicians were less likely to diagnose COPD in women even when spirometry supported this diagnosis.

## Introduction

Women are more likely than men to report delays in the diagnosis of chronic obstructive pulmonary disease (COPD).^1^ However, the etiology of these delays is unknown. Vignette-based studies, involving hypothetical patients who differ only by gender, have found that while physicians are less likely to consider COPD as an initial diagnosis in women, this difference resolves with the performance of spirometry.^2,3^ The implication is that delays in the diagnosis of COPD in women are due to delays in the performance of spirometry. If this hypothesis were true, increasing access to spirometry would yield diagnostic equity in men and women. However, this hypothesis has not been tested with clinical data. We sought to test the alternate hypothesis—that gender differences in the diagnosis of COPD persist after the performance of spirometry—by comparing the time to diagnosis of COPD in men and women with spirometric evidence of obstruction.

## Methods

We used the Optum Labs Data Warehouse (OLDW)—a database consisting of de-identified administrative claims and electronic health record data from across the United States—to identify patients 18 years of age and older with a post-bronchodilator forced expiratory volume in 1 second (FEV_1_) to forced vital capacity (FVC) ratio of less than 0.7 on spirometry performed between 2007 and 2020.^4^ A clinical encounter with a diagnosis code for COPD— including chronic bronchitis and emphysema—was interpreted as a diagnosis of COPD. We excluded patients who had been diagnosed with COPD prior to the performance of spirometry. To better measure the relationship between spirometry and the diagnosis of COPD, we also excluded patients with a positive bronchodilator response, who are more likely to be diagnosed with asthma.

We used a cause-specific Cox proportional hazards model to estimate the time to diagnosis of COPD, accounting for death as a competing risk. The primary exposure was patientc gender, with age, race, ethnicity, tobacco use, and post-bronchodilator FEV_1_/FVC included as covariates.

As the most recent Global Initiative for Chronic Obstructive Lung Disease (GOLD) guidelines recommend repeating spirometry, prior to the diagnosis of COPD, in patients with a post-bronchodilator FEV_1_/FVC *≥* 0.6, in a sensitivity analysis we further compared the time to diagnosis in patients with an FEV_1_/FVC *<* 0.6.^5^

## Results

OLDW included post-bronchodilator spirometry measurements for 4 193 patients, of whom 906 had a post-bronchodilator FEV_1_/FVC *<* 0.7. Of these patients 863 were without a prior diagnosis of COPD, and of the remaining patients, 565 were without a post-bronchodilator response. This cohort consisted of 292 men (52.0%) and 273 women (48.0%) (**Table 1**). The median age was 56 years (interquartile range [IQR] 22). A history of tobacco use was documented for 380 patients (67.6%). The median post-bronchodilator FEV_1_/FVC was 0.64 (IQR 0.10). The median follow up time after spirometry was 24 months (IQR 33). COPD was diagnosed in 200 (35.4%) patients and 20 patients (3.5%) died without a diagnosis of COPD, while 345 patients (61.1%) were right censored.

**Table 1:** Patient Characteristics^*\**^.

|  | **Men**<br>(*n* = 292) | **Women**<br>(*n* = 273) |
| --- | --- | --- |
| **Age, years** | 57 (19) | 55 (23) |
| **Race** |  |  |
| Asian | < 11 | < 11 |
| Black | 27 (9.2) | 25 (9.2) |
| White | 245 (83.9) | 220 (80.6) |
| Other | > 11 | > 11 |
| **Ethnicity** |  |  |
| Hispanic | 27 (9.2) | 23 (8.4) |
| Non-Hispanic | 265 (90.8) | 250 (91.6) |
| **History of Tobacco Use** |  |  |
| Yes | 206 (70.5) | 174 (63.7) |
| No | 86 (29.5) | 99 (36.3) |
| **FEV<sub>1</sub>/FVC** | 0.64 (0.11) | 0.63 (0.10) |
| **Follow Up, months** | 22 (31) | 26 (33) |
| **Diagnosis After Spirometry** |  |  |
| Asthma | 130 (44.5) | 139 (50.9) |
| Bronchiectasis | < 11 | < 11 |
| Chronic Obstructive Pulmonary Disease | 121 (41.4) | 79 (28.9) |
| Cystic Fibrosis | 0 (0) | 0 (0) |
| Any of the Above | 222 (76.0) | 190 (69.6) |
| None of the Above | 70 (24.0) | 83 (30.4) |
| **Treatment After Spirometry** |  |  |
| Short-Acting $\beta_2$ Agonist | 160 (54.8) | 149 (54.6) |
| Long-Acting $\beta_2$ Agonist | 121 (41.4) | 123 (45.1) |
| Long-Acting Muscarinic Antagonist | 43 (14.7) | 34 (12.5) |
| Inhaled Corticosteroid | 146 (50.0) | 146 (53.5) |

\* Values are medians (IQR) for continuous variables and counts (percentage) for categorical variables.
Definition of abbreviations: FEV<sub>1</sub> = forced expiratory volume in 1 second; FVC = forced vital capacity; IQR = interquartile range.

The probability of receiving a COPD diagnosis after the performance of spirometry was lower among women than men (adjusted hazard ratio [aHR] 0.66, 95% confidence interval [CI] 0.50 to 0.88) (**Figure 1**). In our model, the mean time to diagnosis after spirometry was greater in women (106.5 months [95% CI 86.2 to 127.7]) than men (89.7 months [95% CI 72.9 to 106.6]). The probability of receiving a COPD diagnosis was higher in older patients (aHR 1.05 per year of age [95% CI 1.03 to 1.06]) and in patients with a history of tobacco use (aHR 3.61 [95% CI 2.35 to 5.55]). The sensitivity analysis among patients with an FEV_1_/FVC *<*0.6 also revealed a lower probability of COPD diagnosis in women than men (aHR 0.61, 95% CI 0.38 to 0.98).

**Figure 1.**
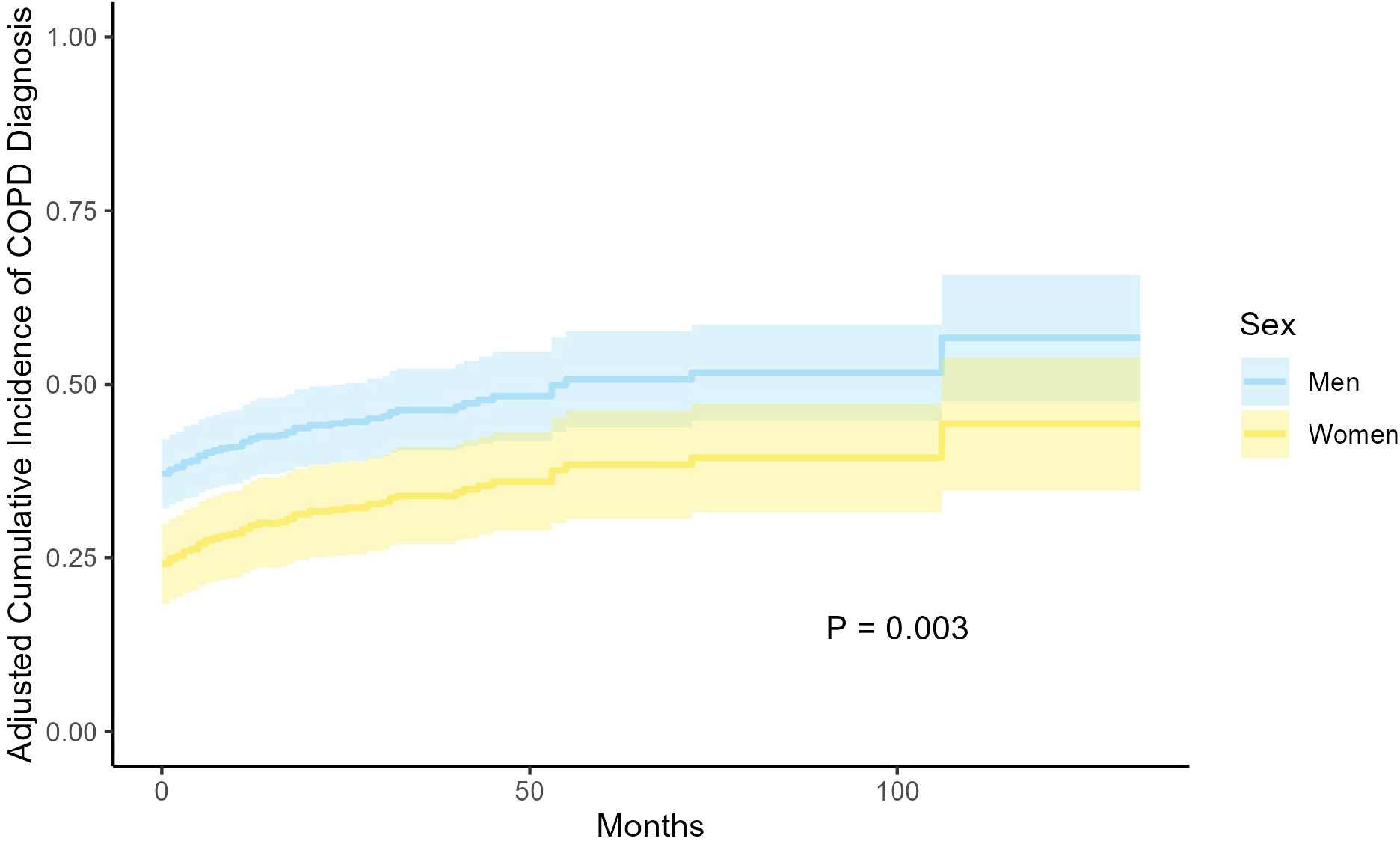
Cumulative incidence function for the diagnosis of chronic obstructive pulmonary disease in men and women, adjusting for age, race, ethnicity, tobacco use, and FEV_1_/FVC. Shaded areas represent 95% confidence intervals. FEV_1_ = forced expiratory volume in 1 second; FVC = forced vital capacity.

## Discussion

In this retrospective cohort study of patients with spirometric evidence of obstruction, the time to diagnosis of COPD was greater among women than men. While previous vignette-based studies have found that gender differences in the diagnosis of COPD resolve with the performance of spirometry, we found that gender differences persist after spirometry has been performed. In our cohort, clinicians were less likely to diagnose COPD in women even when spirometry supported this diagnosis.

Despite the presence of spirometric evidence supporting the diagnosis of COPD, less than half of the patients in our cohort received this diagnosis at any point during follow up, while one fourth of the patients did not receive any diagnosis with which to explain their documented obstruction. More patients were diagnosed with asthma than with COPD and the preference of physicians for this diagnosis—in a cohort defined by the presence of a post-bronchodilator FEV_1_/FVC *<* 0.7 and the absence of a bronchodilator response— is unexpected, particularly as asthma and COPD are not mutually exclusive diagnoses.^6^ While previous studies involving the performance of spirometry have also found low rates of obstructive lung disease diagnosis as well as a preference for the diagnosis of asthma over COPD, our study is the first to suggest that this preference is present even in patients for whom spirometry has already been performed.^7,8^ Our study suggests that the underdiagnosis of COPD stems from more than just the underuse of spirometry.^9,10^

Our study has several important limitations. First, though spirometric evidence of obstruction is necessary for the diagnosis of COPD, it is not sufficient, with the diagnosis dependent as well upon the presence of appropriate symptoms or risk factors.^5^ Data regarding the presence of symptoms and risk factors are not included in the OLDW and gender differences in symptoms and risk factors may account for some proportion of the diagnostic differences identified in this study. Second, though our cohort was drawn from a national database, the inclusion of PFT data in this database was subject to selection bias, potentially limiting the generalizability of our findings.

In summary, in a retrospective cohort of patients with spirometric evidence of obstruction, the time to diagnosis of COPD was greater among women than men. Further work is needed to understand the etiology of this difference.

## Data Availability

All data produced in the present study are available upon reasonable request to the authors

